# Colibactin-associated mutations in the human colon appear to reflect anatomy and early exposure, not oncogenesis

**DOI:** 10.64898/2026.04.13.26350783

**Authors:** Laurel Hiatt, Elizabeth V. Peterson, Hannah C. Happ, Joshua Major-Mincer, Akshay Avvaru, Camila L. Goclowski, Alexis Garretson, Thomas A. Sasani, James M. Hotaling, Deborah W. Neklason, Amiko M. Uchida, Aaron R. Quinlan

## Abstract

Colorectal cancer (CRC) is the second leading cause of cancer death globally and the number one cause of cancer death in people under 50 years old. The reasons for the rise of early-onset CRC are unknown, and while anatomically distinct subtypes of CRC have substantial clinical and molecular associations, the etiology of region-specific disease, such as early-onset CRC’s enrichment in the distal colon, remains unclear. Understanding regional mutagenesis may identify risk factors for this public health concern and CRC more broadly. To evaluate mutational dynamics across the premalignant colon, we performed whole-genome sequencing of 125 individual colon crypts taken from six standardized regions biopsied during colonoscopy, collected from 11 donors without polyps and 10 with polyps. We observed mutation spectra and accumulation rates consistent with previous whole-organ studies, with greater subclonal mutation capture enabled by experimental design. T>[A,C,G] mutations, which are associated with colibactin genotoxicity from pks+ *Escherichia coli*, were significantly enriched in the rectum of donors with and without polyps (adjusted p-values < 0.01). Moreover, when comparing findings to crypts from individuals with CRC and sequenced CRC tumors, we observed consistent enrichment of the colibactin-associated mutational signature “ID18” in the rectum in both normal colon crypts and CRC tumors, without significant difference in colibactin-specific single nucleotide variant or insertion-deletion burden in crypts across the three clinical groups (i.e., no polyp, polyp, and CRC). These findings argue against a causal or prognostic role for colibactin in CRC, instead indicating that the proposed association with early-onset disease reflects anatomic specificity rather than cancer-specific clinical relevance.

## Introduction

Colorectal cancer (CRC) is the third most common cancer and the second leading cause of cancer death globally, with incidence predicted to increase by 60% in developing countries by 2030.^1^ Although CRC mortality has dropped over the past several decades in those older than 50, incidence and mortality have risen in younger patients, who often have worse prognoses in part due to delayed detection.^2,3^ From 2004 to 2014, early-onset (i.e., before age 50) CRC mortality rose *annually* by 0.9–1.9%, depending on decade of life.^4^ There have been repeated calls to lower the initial age recommended for colonoscopy screening,^5^ but limitations to clinical implementation remain due to uncertainties about disease progression.^6,7^ Without tools to determine patient-specific prognoses from premalignant colonoscopic findings, we cannot adjust screening regimens to address clinical needs. Instead, we risk untenable clinical burdens for both clinical workforces and patients, resource waste,^8^ and, in rare cases, patient harm.

It is therefore crucial to understand the pathogenic processes underlying early-onset CRC to better inform preventive and prognostic clinical efforts.^9^ Recent clinical research has shown that sporadic early-onset cases in the United States are most common in the distal or “left” colon.^10^ Similarly, a global overview of colorectal cancer found early-onset CRC to be 1.88-fold enriched in the distal colon/rectum.^11^ The rising incidence of left-sided CRC in early-onset cases is speculated to reflect evolving environmental factors associated with industrialization, yet the etiology of regional disease remains unclear.^12^ This knowledge gap is particularly significant given the well-established regionalization of CRC: tumor location has been used as a surrogate for various pathologic attributes, such as neoplastic morphology and clinical prognosis.^13–15^ CRC subtypes also differ by genetic factors, such as driver mutations and microsatellite instability,^16^ which in turn are strongly associated with anatomical site.^17–19^ Therefore, understanding regional mutagenesis in the context of regional disease presentation could clarify CRC etiology, particularly for early-onset disease, where distal colon enrichment remains mechanistically unexplained.

Studying somatic mosaicism in healthy or pre-disease tissue is proposed to improve understanding of disease development.^20,21^ Evaluating premalignant colon is especially promising given the step-wise tumorigenesis observed in CRC: although exact mutational thresholds and biological transitions are still being characterized,^22,23^ CRC is known to progress in a step-wise fashion. Colon crypts, ∼2000-cell structures comprising the colon’s absorptive epithelium, become dysregulated and form adenomatous polyps,^24^ which confer disease risk and can progress to cancer over time if left untreated.^25^ While colonoscopies facilitate adenomatous polyp discovery and removal—with future screening informed by polyp size and morphology^26^—there is increasing evidence that even “normal” tissues harbor mutations associated with eventual disease.^27,28^ If we can understand which mutations predispose a tissue for carcinogenesis and which are tolerated long-term, we can design clinical strategies that are more sensitive to disease risk and capable of personalized prognoses, thereby reducing the clinical and economic burden of current screening uncertainty. However, previous work evaluating the mutational landscape of the premalignant colon has been performed at the organ level,^6,7,29^ despite the clinical significance of anatomic regions in CRC. Determining regional patterns of mutagenesis in premalignant colon may reveal mutational etiologies underlying regional CRC, which, in turn, could elucidate factors involved in distally enriched cases of early-onset CRC.

Notably, the bacterial product colibactin has been proposed to be causative and potentially prognostic in CRC, especially early-onset.^30–32^ Mutational signatures are inferred patterns of sequence-specific mutations associated with assorted mutagenic events or sources,^33^ and the colibactin mutational signature, ID18, is enriched in left/rectal CRC.^17^ As colibactin mutagenesis patterns share regional enrichment with early-onset CRC, evaluating mutational landscapes in premalignant colon may clarify hypotheses of colibactin’s causal role and predictive potential. Furthermore, establishing a biological baseline is essential to defining the threshold between a healthy colon and incipient cancer, as the initiation of tumorigenesis is still largely uncharacterized.^34^

In partnership with the University of Utah’s Microbiota and GI Immunology Consortia (MAGIC) Biobank, we assessed mutational landscapes of the premalignant colon across six anatomic sites in 21 patients. The cohort comprises 11 patients with no polyps identified at colonoscopy and 10 with at least one adenomatous polyp. Using laser capture microdissection, we isolated individual colon crypts, which are monoclonal and therefore genetically homogeneous due to their contained microstructure and confined stem cell niche.^35^ We quantified baseline mutagenesis across discrete anatomic sites of the colon to evaluate whether mutational shifts differ by region or polyp status in the premalignant colon. We incorporated two sources of previously published data into our analyses: 1) colon crypts from individuals with CRC (n=15)^29^ and 2) mutational trends from a regionally annotated study of CRC tumors^17^. The latter study evaluated six mutational signatures that overlap with those we extract (summarized trends and known associations in **Supp. Table 1**). By comparing premalignant colon mutagenesis to tumor-derived mutational trends, we identify mutational patterns shared between premalignant tissue and established tumors—including the closely comparable patterns of signature ID18 burden across clinical groups indicative of common colibactin mutagenesis—and identify divergences that may reflect disease-specific processes.

## Results

### Study overview and overall mutational landscape

We conducted whole-genome sequencing of 125 individual crypts isolated by laser-capture microdissection from the premalignant colon of 21 individuals: 11 without colonoscopy findings and 10 with polyps (**Fig. 1A**). Biopsies were taken from six standardized regions during colonoscopy, in the following order: cecum, ascending colon, transverse colon, descending colon, sigmoid colon, and rectum (see **Methods**). These regions can also be grouped as the right (cecum, ascending, transverse) and left (descending, sigmoid, and rectum) colon. A median of five regions per donor were represented in our data, with the number of individual crypts per donor ranging from 2 to 15 (mean and median: 6; **Table 1**). We were able to achieve high DNA sequencing quality (average sample base quality at or above Q30: 93.91%) and median genome-wide DNA sequencing coverage of ∼26X (**Supp. Fig. 1**). Since colon crypts are clonal structures arising from a few stem cells, this sequencing coverage facilitated the accurate detection of both clonal and subclonal mutations, defined by a mutation allele frequency (MAF) ≥ and < 0.2, respectively (**Fig 1B**).

**Figure 1:**
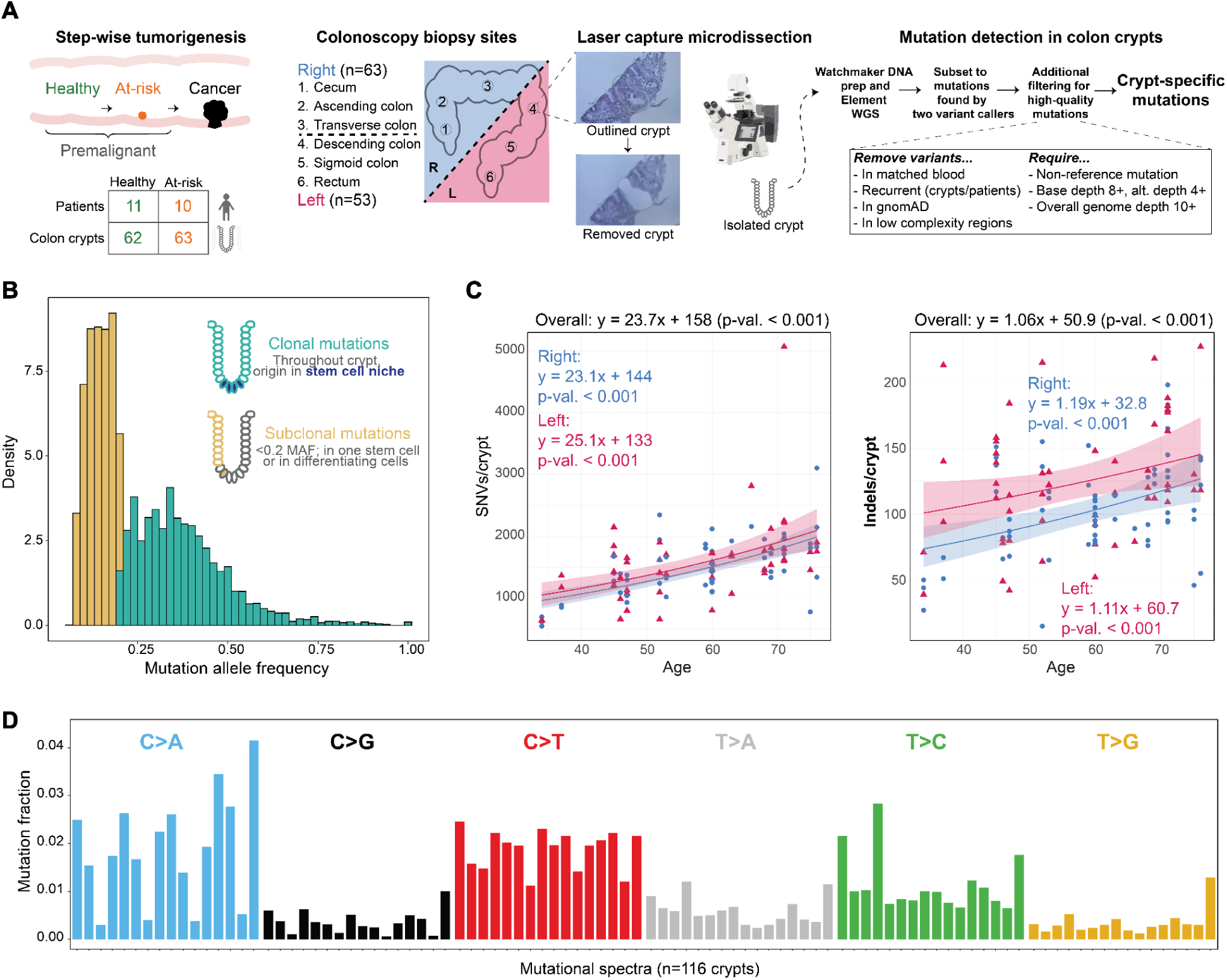
Study overview: experimental design and data generation. **A.** Experimental design of study; step-wise tumorigenesis is shown as a schematic corresponding to patient and crypt counts across the two premalignant groups. Biopsy sites are shown from right (R) and left (L), and a schematic with associated pictures of laser capture microdissection work flow is shown in addition to data generation and filtering strategies. **B.** Mutation allele frequency (MAF) for all mutations passing filtering thresholds in 125 crypts from study. Distribution of clonal (≥ 0.2) and subclonal (<0.2) MAF outlined in teal and yellow respectively, with explanatory schematic of colon crypt dynamics. **C.** Crypt-specific (n= 116, high-confidence samples, see **Methods**) mutations for SNVs and indels shown as a negative binomial GLM (see **Methods**) of burden over age to show accumulation over time; parsed by right (n=63, magenta triangles) and left (n=53, blue circles). The darker gray around the lines is the 95% confidence interval. **D.** The mutational proportions, by trinucleotide context, of the 116 high-confidence crypts.

**Table 1:** Patient demographics. All patients are male. Two patients without polyps (italicized) did not have matched blood samples. Polyp size is classified as small (<5mm), medium (5-9mm), and large (10+ mm). Biopsy abbreviations are as follows: cecum (C), ascending colon (A), transverse colon (T), descending colon (D), sigmoid colon (S), and rectum (R), with the initial number indicating the total number of sites.

| **Polyp(s)** | **Age** | **Race** | **Biopsies: regions** | **Indication for endoscopy** |
| --- | --- | --- | --- | --- |
| None | 30's | White | 5: C, A, T, D, R | Surveillance: genetic risk |
| None | 30's | White | 6: C, A, T, D, S, R | Screening: gastrointestinal symptoms |
| None | 40's | *Hispanic/Latino* | 6: C, A, T, D, S, R | *Screening* |
| None | 40's | White | 4: A, D, S, R | Surveillance: previous polyps |
| None | 40's | Asian | 5: C, T, D, S, R | Screening |
| None | 50's | White | 9: C, A, T, T, D, D, S, S, R | Surveillance: previous polyps |
| None | 50's | White | 5: C, A, T, D, R | Screening: gastrointestinal symptoms |
| None | 50's | White | 3: C, A, T | Surveillance: previous polyps |
| None | 60's | White | 5: C, T, D, S, R | Surveillance: previous polyps |
| None | 70's | *White* | 8: C, A, T, D, S, S, R, R | *Surveillance: previous polyps* |
| None | 70's | White | 6: C, A, T, D, S, R | Screening |
| 2 small, distal | 40's | White | 5: A, T, D, S, R | Screening |
| 4 small/medium, transverse and distal | 40's | White | 6: C, A, T, D, S, R | Screening |
| 2 small, distal | 60's | White | 5: C, A, T, D, R | Surveillance: previous polyps |
| 2 small/medium, proximal and distal | 60's | White | 15: 7C, 3A, T, T, D, S, R | Surveillance: previous polyps |
| 2 small, proximal | 60's | Black | 2: T, S | Surveillance: previous polyps and gastrointestinal symptoms |
| 3 small/large, transverse | 60's | Hispanic/Latino | 7: C, A, T, T, D, R, R | Surveillance: previous polyps |
| 4 medium, proximal and distal | 60's | White | 6: C, A, T, D, R, R | Surveillance: previous polyps |
| 3 medium/large, proximal | 70's | White | 5: C, A, T, D, R | Surveillance: previous polyps |
| 2 small/medium, proximal and distal | 70's | White | 6: C, A, T, D, S, R | Surveillance: previous polyps |
| 1 large, proximal | 70's | White | 6: C, A, A, T, S, R | Referral |

As expected and documented in prior studies, the number of somatic mutations observed per crypt accumulated significantly with age (**Fig. 1C**).^36,37^ We found an overall annual accumulation of ∼23.7 SNVs per year (range ∼20–40; **Supp. Fig. 2**). This estimate is comparable to a previous study estimate of ∼36/year, with a 95% confidence interval of ∼26.9 to 50.6.^38^ The overall accumulation of ∼24 SNVs/year was consistent across the right and left colon, and the indel accumulation rate was also consistent by side (∼1/year). Furthermore, we compared burden ranges to those reported previously in 50 crypts from seven individuals with a similar age range (this study: 34–76 years; Moore^39^: 38–78). SNV burden ranges (this study: 545–5066; Moore: 883–5160) and indel burden ranges (this study: 15–227; Moore: 20–147) were both comparable. We may observe a higher maximum burden due to our sequencing technology’s increased sensitivity toward indels^40^ or individual-specific biology. The most abundant mutational type was C>T, which accounted for >30% of mutations, and the overall mutation spectra (i.e., frequency distribution of different mutation types such as C>T, C>A, etc.) were consistent with mutation spectra observed in previous studies of crypt mutagenesis (**Fig. 1D**),^29,41^ indicative of shared tissue-specific biology. When comparing mutational spectra to those of healthy males from the Lee-Six et al. study (a similar biological context), the cosine similarity is 0.981, indicating comparable mutagenesis of these “normal” crypts (**Supp. Fig. 1**).

### Region-specific mutational dynamics

We next examined variation in mutation type and burden across the six biopsied regions to identify regional differences in premalignant colon mutagenesis. We observed consistent microsatellite stability across regions in premalignant samples as determined by a panel of five diagnostic loci (**Fig 2A**, see **Methods**). We used the statistical method ‘aggregate mutation spectrum distance’^42,43^ to compare the mutation spectra of crypts from the left and right colon. By evaluating the spectra distance with 10,000 simulations permuted from SNV trinucleotide context matrices, we found that the right and left colon differed significantly by mutation proportion (p-value = 0.008) but not by mutation count (p-value = 0.119) (**Fig. 2B**), indicating that while overall SNV counts are comparable across sides, the composition of mutation subtypes differs.

**Figure 2:**
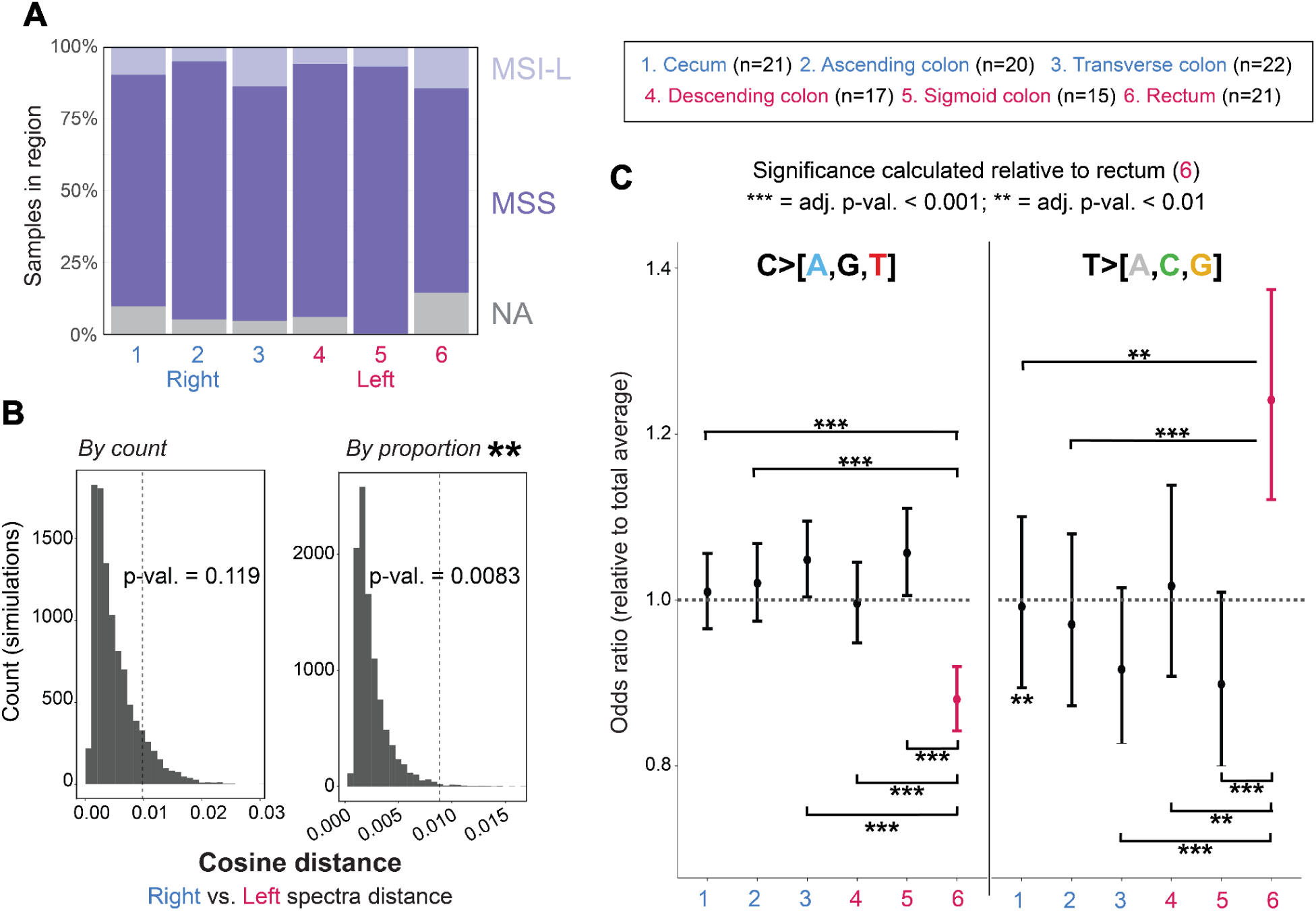
Premalignant colon varies regionally by specific mutation composition. **A.** Microsatellite instability across six regions colored by side shown as either NA (gray; insufficient evidence), MSS (dark purple, microsatellite stable), or MSI-L (microsatellite-instability low, light purple). Region sample counts (also in C) are: cecum (n=21), asc. col. (n=20), trans. col. (n=22), desc. col. (n=17), sig. col. (n=15), rectum (n=21). **B.** Aggregate mutation spectra distance shown for right and left samples in 96-trinucleotide context with by count (“sum” argument) and proportion (“mean” argument). **C.** Mutation subtype, collapsed to C>[A,G,T] and T>[A,C,G], are shown relative to the overall average prevalence as an odds ratio with a confidence interval; regional significance is calculated as subtype proportion relative to the rectum (point and bar shown in pink), as the only region entirely shifted from the total average for both mutation groups (see **Methods.** P-values adjusted by Benjamini-Hochberg.

When modeling mutation burden adjusted for age and clinical group (**Methods**), SNV burden did not differ significantly by side (right mean: 1517, interquartile range: 546; left mean: 1563, IQR: 546). However, when evaluating SNV composition across the colon, we found that the C>T subtype was significantly enriched on the right side (adj. p-val. = 0.026, paired Wilcoxon tests). Additionally, C>A, C>G, T>A, T>C, and T>G substitutions were significantly different when comparing across regions, with the strongest signal in the T>A and T>C subtypes (adj. p-values < 0.05, Kruskal–Wallis tests) (**Supp. Table 2**). When comparing to the global average for mutations grouped into C>[A,G,T] and T>[A,C,G], only the rectum was entirely distinct from the global average (**Fig. 2C**), and was significantly different from each region with regard to both mutation groups. T>[A,C,G] mutations were significantly elevated in the rectum compared to the global mutation average and to other regions, while C>[A,G,T] were comparatively depleted.

Similarly, indel mutation burden was significantly increased in the left colon (mean: 124, IQR: 54) versus right (mean: 103, IQR: 47) (adj. p-val. < 0.001), with an approximate increase of ∼26% in the model (**Methods**).

### Mutational signatures across clinical groups

In light of the developing hypothesis that the genotoxin colibactin is causative of CRC, we made note of the previously-observed association of colibactin mutagenesis with indels^44^ and T>[A,C,G] mutations.^45,46^ As we observe an impact of colon anatomy on indel and T>[A,C,G] mutation abundance, we hypothesized that these variations may reflect regional mutagenic processes, which in turn should be observable as mutational signatures. We extracted signatures from the premalignant samples and found the two signatures associated with genotoxic colibactin to be significantly enriched in the rectum: single base substitution signature SBS88 and indel signature ID18^30^ (**Fig. 3A, Supp. Fig. 3**). To distinguish region-specific from disease-state-specific mutagenesis, we compared these findings to regional tumor trends from Cornish et al. **(Supp. Table 3)**, which identified ID18 as increasing from right to left in colorectal tumor-bearing patients (**Supp. Table 1**). We separated the premalignant dataset into patients with normal colonoscopic findings (’healthy,’ n=11) and those with adenomatous polyps (’at-risk,’ n=10), and compared both groups to the tumor trends, scaling results to overlay the trend lines (**Fig. 3B**).

**Figure 3:**
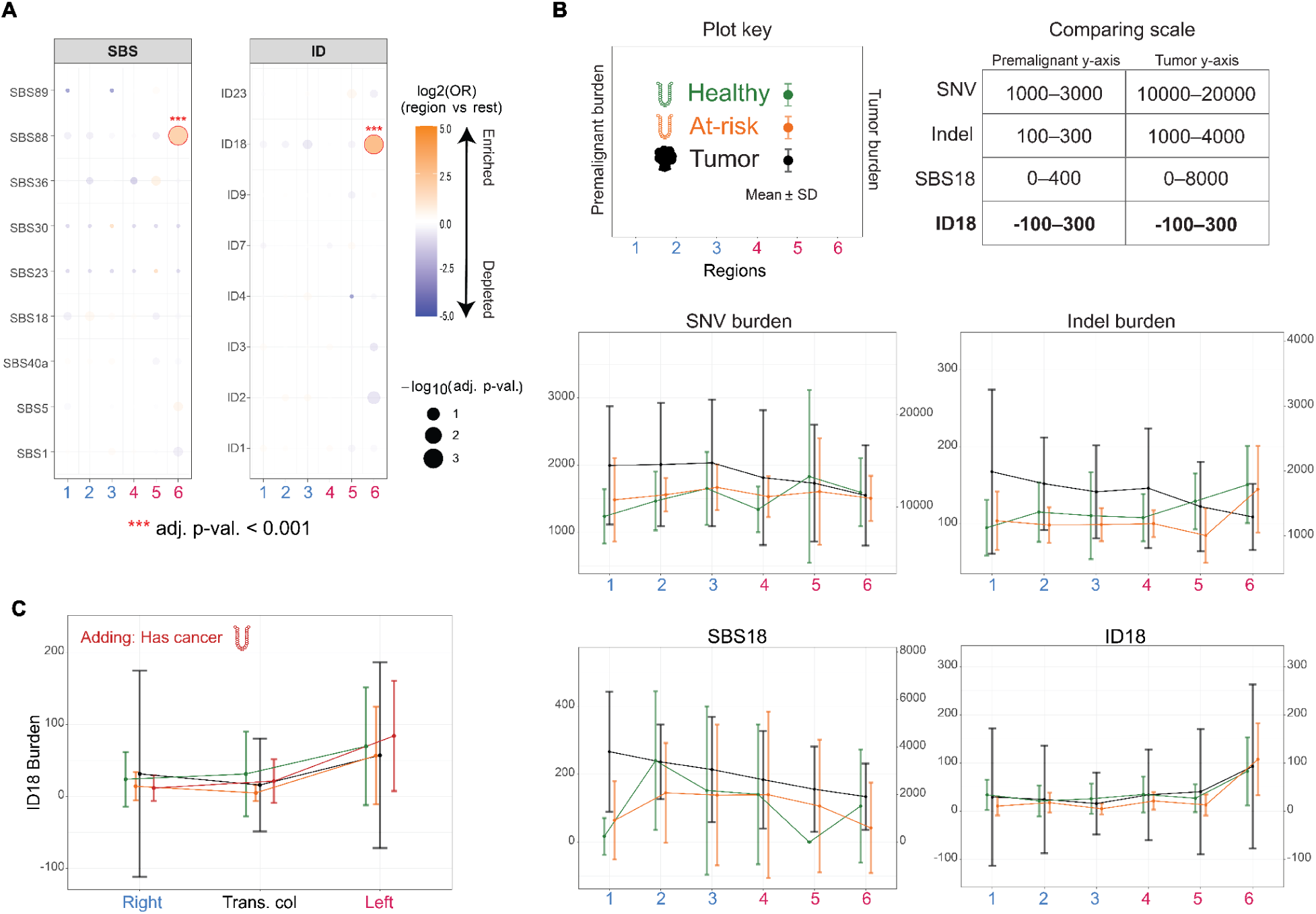
The rectum shows colibactin enrichment regardless of clinical status. **A.** Regional enrichment (orange) and depletion (purple) for each non-artifactual signature in this study derived from odds ratios as determined by logistic regressions, adjusted for age, clinical group, and random patient effects (**Methods**). P-values adjusted by Benjamini-Hochberg. **B.** SNV and indel burdens, as well as SBS18 and ID18 burdens, compared between clinical groups, integrating publicly available tumor data from Cornish et al. (2024) at overlapping regions with this study’s premalignant data. The tumor data has a separate burden axis scaled to match the premalignant visualization. Plot key for interpretation of plots is shown at bottom of panel, as well as a table of y-axis comparisons, highlighting the consistent burden of ID18 between clinical groups. Premalignant data include 59 healthy:57 at-risk samples, subset by region as 10:11 (1: cecum), 9:11 (2: ascending colon), 10:12 (3: transverse colon), 10:7 (4: descending colon), 9:6 (5: sigmoid colon), and 11:10 (6: rectum). Healthy: green, at-risk: orange, Cornish (tumor) trend: black. Mean and standard deviation shown for all. All SBS signature comparisons shown in **Supp. Fig. 4**, and all ID signature comparisons shown in **Supp. Fig. 5**. C. ID18 is evaluated with the addition of Lee-Six crypts from individuals with and without CRC, compared as burden means with standard deviation across the three annotated regions—right, transverse colon, and left—combined with premalignant data and compared to the the Cornish tumors (black). *Healthy* (green): right n=100, transverse colon n=76, left n=71; *at-risk* (orange): right n=22, transverse colon=12, left n=23; *has cancer* (red): right n=35, transverse colon n=35, left n=43.

We stratified mutations by clonal (≥0.2 MAF) and subclonal (<0.2 MAF) status to distinguish signatures clonally established in stem cells from those restricted to a subset of crypt cells (**Supp. Fig. 4; Supp. Fig. 5**). Patterns of signature burden subset to clonal mutations largely recapitulated overall trends given their higher contribution to total counts. Premalignant groups were discordant with tumor trends in SNV and indel burden (**Fig. 3B**); this discordance was expected given the absence of microsatellite instability in these samples, which likely drives elevated proximal burdens in tumors.

Although lower in count and thus contribution to the overall signature trends, subclonal mutations still revealed distinct patterns. The slippage signature ID2 was proximally enriched in at-risk and tumor samples in the overall data, yet the increased concordance in the subclonal data—including the apparent proximal enrichment in health samples— suggests this signature may be increased proximally in specific mutagenetic contexts (**Supp. Fig. 5**). This subclonal pattern might represent unrepaired mutations in differentiating cells or mutations restricted to a stem cell subset. Given that ID2 is enriched in proximal tumors, subclonal slippage damage may be resolved in normal stem cells but persist in dysregulated cancer cells. Similarly, the at-risk group aligned with tumor trends for subclonal SNV, indel, and SBS5, potentially reflecting the older average age (the patient group with polyps is 10 years older on average) of this group or an advanced clinical state. All three clinical groups were concordant for subclonal SBS1, which—as with ID2—may reflect unrepaired damage in differentiating cells or damage to a subset of the stem cell niche.

Notably, two regional trends were concordant across all three clinical groups: a proximal-to-distal decrease in burden for signature SBS18, and a proximal-to-distal increase in burden for ID18. Both signatures were absent from the subclonal dataset, suggesting either early mutagenic exposure and/or long-term persistence of damage in stem cells. Critically, ID18 — the *E. coli* colibactin-associated signature — showed a consistent regional pattern of comparable mutational burden across all three clinical groups, with no rescaling required to overlay premalignant and tumor data (1:1 scale; **Figure 3B**). This is in direct contrast to all other signatures examined, in which the tumor dataset has substantially elevated mutation burdens requiring an adjusted visualization scale (range 1:5 to 1:250)—underscoring that ID18 burden in premalignant tissue was indistinguishable from that in tumors.

The preceding comparison evaluated ID18 in premalignant crypts against tumor-derived mutations, which differ in tissue context and genotyping approach. To enable a more direct comparison, we combined these data with isolated crypts from Lee-Six et al — including crypts from patients with CRC — genotyped through our pipeline (**Methods**). This yielded a dataset of crypts from healthy, at-risk, and cancer patients across three regional contexts (right, left, and transverse colon). Even with this expanded, methodologically consistent comparison to reveal more granular differences, ID18 burden remained consistent across all three clinical groups, despite hypotheses predicting elevated colibactin mutagenesis in individuals who develop CRC (**Fig. 3C**).

### Colibactin-associated mutational dynamics

Recent work has proposed that colibactin as a genotoxin leads to subsequent CRC.^47–52^ In this case, it would be expected that ID18 mutations would vary across clinical groups and that healthy colon crypts would have fewer ID18 mutations than malignant tissues. However, the analysis incorporating Cornish et al. and Lee-Six et al. data show consistency in ID18 mutations independent of clinical status. Instead, ID18 varies by anatomic site, with distal colonic rectal tissue bearing the highest burden of these mutations. Additionally, the colibactin-associated SBS88 signature^30^ is similarly consistent across clinical groups and regionally enriched in the rectum (**Supp. Fig. 6**; **Fig. 3A**).

To more directly identify colibactin-derived mutations, we evaluated SNVs and indels across all three clinical groups at sequence motifs with demonstrated colibactin mutagenesis (**Methods**).^53^ These motifs reflect colibactin’s preferential binding to AT-rich sequences.^54,55^ Briefly, a previous study leveraged colibactin-exposed organoids to detect and identify sequence-specific motifs preferentially mutated by colibactin, in a 10bp window extended from mutations. We applied these extended-context motifs to identify colibactin-associated SNVs and indels in the aggregate dataset (**Fig. 4A**), reasoning that motif-level analysis might detect differences across clinical groups or age that the inference-based signature extraction process might miss.

**Figure 4:**
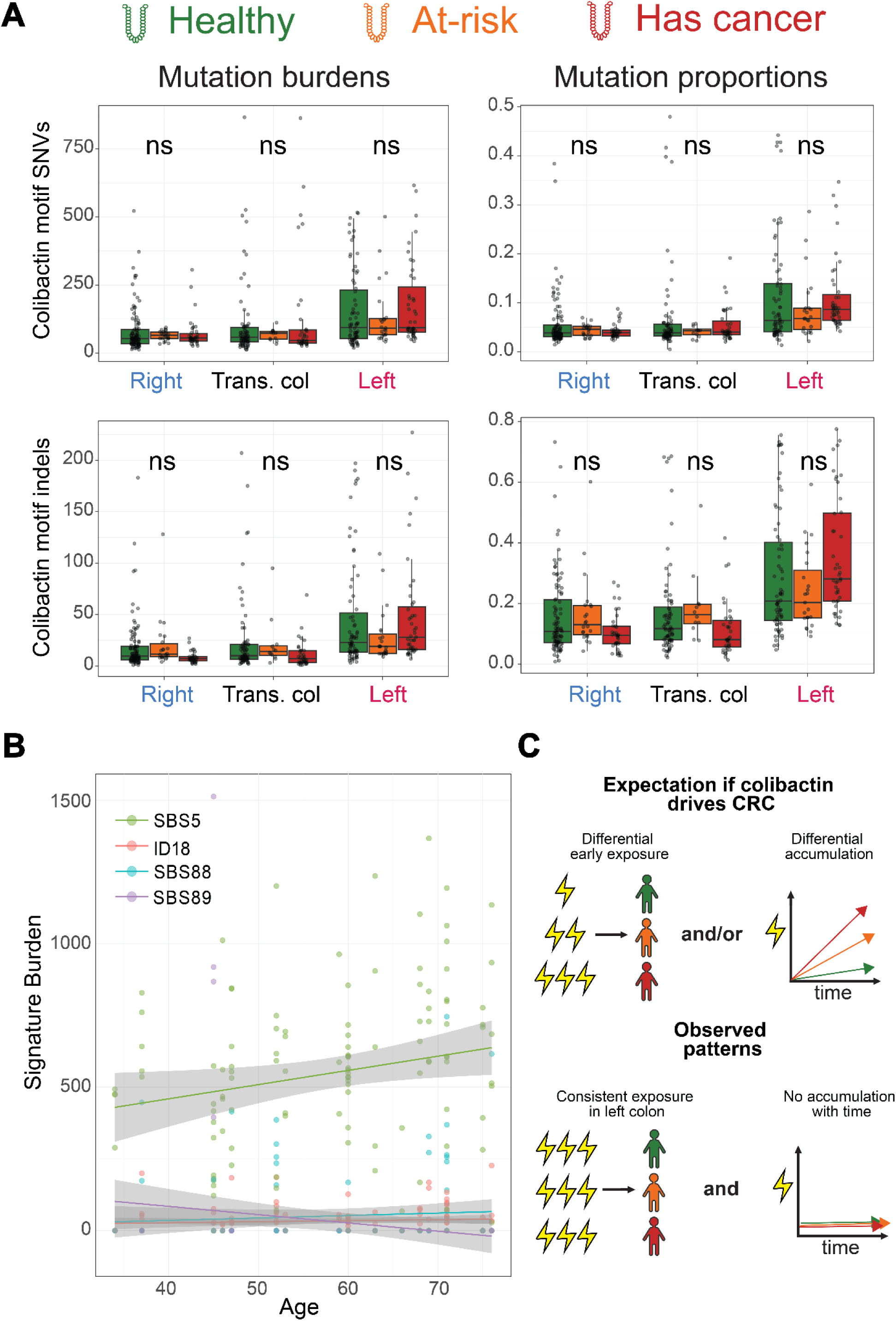
Colibactin-associated mutations are consistent in states of health and malignancy. Premalignant data is derived from this and the Lee-Six study, while the “has cancer” data derives from the normal crypts of CRC patients in the Lee-Six study. **A.** Mutation burdens and proportions across regions for SNVs and indels matching colibactin motif sequences. Significance between clinical groups determined by GLM modeling with Benjamini-Hochberg correction (**Methods**); ns: not significant. **B.** Linear regressions of a clock-like signature (SBS5) and a signature associated with early life exposure (SBS89), framing the flat lines of colibactin signatures SBS88 and ID18. Gray shading indicates 95% confidence interval. **C.** Schematic comparing potential models for colibactin initiating or predisposing carcinogenesis, compared to the model supported by the data.

However, colibactin-associated SNV and indel counts and proportions were still comparable across the three clinical groups (all adj. p-values > 0.05). Stratifying by genic and coding regions did not change the overall consistency across clinical groups, although power for this analysis was limited (the median number of colibactin SNVs within an exon was 0 per genotyped sample) (**Supp. Fig. 7**). Colibactin-associated SNVs showed high cosine similarity to SBS88 (0.96 in healthy, 0.93 in at-risk), indicating accurate motif-based detection consistent with signature extraction (**Supp. Fig. 8**). When we modeled colibactin-associated SNV and indel burden and proportion using generalized linear mixed models including age, the only significant differences were regional (i.e., enrichment in distal colon), not between clinical groups. Age was not a significant model factor for colibactin mutation burdens, suggesting that these are early acquired mutations rather than accumulated over the lifetime. Moreover, colibactin signatures track the age trajectory of SBS89, a signature understood to be acquired early in life,^11,17^ rather than a clock-like signature such as SBS5 (**Fig. 4B**). Together, these results indicate comparable early-life colibactin exposure across clinical groups, with no evidence of differential initial burden or ongoing accumulation that would be expected if colibactin mutagenesis were an initiating factor in CRC (**Fig. 4C**). If colibactin mutagenesis was a driving disease factor, we would expect to see a higher initial burden in individuals with an early life exposure, or increased accumulation over time; instead, we see data consistent with an early life exposure that is comparable across clinical groups, regardless of disease status or age.

## Discussion

Colibactin-associated mutagenesis in the colon is site-specific and independent of disease state. These mutations appear to result from early life exposure and persist without innate pathogenicity, contradicting current theories that colibactin-driven mutagenesis predisposes individuals to CRC. A previous study identifying colibactin signatures in early-onset CRC cases proposed that examination of normal crypts across groups of variable clinical states should reveal elevated SBS88 and ID18 if colibactin mutagenesis causally drives early-onset CRC.^11^ However, our data are contrary to this hypothesis as we found SBS88 and ID18 levels are comparable across patient age and clinical status and appear to reflect early-life, region-specific exposure rather than ongoing mutagenesis. Interestingly, the ID18 burden reported by this previous study in microsatellite stable colorectal tumors (which are 76% distal/rectal) falls well within the confidence intervals we observe for left colon crypts across the data groups (mean 110, median 111). However, the authors of Díaz-Gay et al.—as is common practice— report signature data in their manuscript as percentages of the individual’s signature burden, which may reflect a broader approach obscuring the clinical actuality of colibactin mutagenesis.

Because colibactin mutations accumulate early in life, younger patients naturally carry a higher *proportion* of these mutations relative to their lower overall mutation burden—making colibactin mutations appear enriched in younger individuals (**Supp. Fig. 9**) without any difference in absolute exposure (**Supp. Fig. 6**). In analyses of colibactin-associated motifs, age was not a significant factor when modeling colibactin mutations by count, whereas it became a significant factor when modeling by proportion. This suggests that while the numerator of colibactin mutations remains consistent across groups, the denominator of total mutations increases over time. Furthermore, the ID18 signature is detected only in clonal expansions and does not accumulate with age, consistent with a limited, early-life event rather than continuous, cumulative colibactin activity.

The elevation of the colibactin signal in the rectum, where sporadic early-onset cancers are especially common, likely exacerbates the conflation of biological incidence with clinical relevance. Although overall mutation counts were consistent across anatomic sites, regional mutational spectra differ, particularly in T>[A,C,G] subtypes associated with colibactin mutagenesis.^45,46^ The data showed that colibactin-associated SNV and indel burden did not differ across clinical groups (p-values > 0.05), with significant differences only by region showing distal enrichment. Indeed, colibactin mutagenesis appears to be a common physiological event that is regionally, rather than clinically, distributed. These findings align with a recent report that more than half of enrolled infants were colonized by a colibactin-producing *E. coli* strain during their first two years of life, leading the authors to conclude that colibactin exposure is part of normal gut microbiome development and unlikely to independently increase cancer risk, given its prevalence far exceeds that CRC in the general population.^56^ The data support this interpretation, which is a pressing consideration given the numerous cases in which colibactin has been proposed as a causative factor in CRC^48,49,57^ and as a potential prognostic factor for early-onset CRC^32,48^.

Instead, a more parsimonious explanation is that sequence-specific colibactin adducts can form within clinically relevant genes as part of stochastic mutagenesis,^58^ which, in turn, leads to apparently recurrent driver mutations matching the colibactin signature, as previously reported^11,31,59^. This follows a recent study hypothesizing that “short linear motifs” mutated by distinct signature-associated mutagenesis are a mechanistic component of cancer genomes, mediating oncogenic transformation through indirect sequence specificity.^60^ As such, interrogating driver mutations in patient mucosa, regardless of colibactin association, may be more informative than inferring clinical significance from aggregate colibactin burden. It is also necessary to distinguish the carriage of pks+ *E. coli,* which colonizes opportunistically in inflamed conditions and appears more abundant in early-stage CRC than in healthy intestinal mucosa or later-stage cancer,^61^ from mutations incurred from genotoxic exposure decades prior. In fact, multiple studies have found no direct relationship between colibactin genes indicative of current intestinal colonization and colibactin-associated mutagenesis, reinforcing this distinction.^47,62^

More generally, our study leveraged technical advances including library preparation optimized for low DNA input and high-fidelity sequencing^40^, to improve sequencing accuracy and overall coverage. This consequently enabled the capture of high-confidence subclonal mutations, which was limited in previous studies.^29,39^ Because multiple descendants of an ancestor stem cell maintain the crypt population, low-frequency mutations may occur through this mechanism or through unrepaired damage in differentiated cells.^24^ Where subclonal mutagenesis differs from clonal and overall trends, such as in SNV/indel burdens and SBS1 by region, it may reflect biology unobserved in past colon crypt studies. These unique aspects of mutagenesis, apparent only in the subclonal mutations of normal crypts, may play a role in tumorigenesis as mutation repair becomes increasingly dysregulated. Conversely, signatures that were consistent across biopsy sets and clonality states may reflect innate organ biology. For example, the oxidative signature SBS18 globally decreased in burden from proximal to distal colon, which may reflect greater oxidative stress in the proximal colon from factors such as increased electrolyte transport and bile acid exposure.^1,15^ This proximal enrichment is consistent with the association of oxidative damage with microsatellite instability and mismatch repair deficiency,^63,64^ both of which are known to be enriched in the proximal colon.^14,65^ However, as SBS18 was also found to be proximally enriched in microsatellite-stable CRC and in this dataset, further clarification is needed to distinguish region-specific phenomena from pathology-specific mutagenesis.

This study has limitations. First, the sample size of each patient population was small, limiting our ability to broadly generalize findings. We sampled from standardized regions and did not biopsy immediately adjacent to a polyp, nor did we sequence polyps. Second, we had distinct methodological approaches from the data sets we aggregate in analyses.^17,29^ Additionally, our data appears potentially enriched for subclonal C>A artifacts likely due to damage during library generation (**Supp. Fig. 10**). Still, as these are not relevant to colibactin signatures or sequence motifs—which persist when subclonal mutations are removed—and were identified as artifacts during signature extraction, it is unlikely to affect central interpretations.

We conclude that distinguishing the carriage of colibactin-producing microbes from colibactin-associated mutations is clinically essential. While colibactin has been a compelling candidate in colorectal carcinogenesis, the data argue that its mutational footprint is regionally determined, early-acquired, and independent of disease state, undermining its proposed role as a causal or prognostic factor in CRC. Further research is essential to disentangle apparently common colibactin mutations, colibactin-producing *E. coli* carriage, and the factors that definitively initiate and progress tumor formation. Careful analysis of specific crypt mutations may be leveraged in the future to determine patient risk and personalized screening regimens. These precision-focused analyses and clinical strategies, in turn, may reduce the clinical morbidity of high-risk CRC, including early-onset cases. Ultimately, the clinical value of crypt mutagenesis may lie not in aggregate signature burden but in the identification of specific driver mutations, whether or not colibactin-associated, that confer individual risk and inform personalized screening.

## Materials and Methods

### Data reporting

No statistical methods were used to predetermine sample size. Experiments were not randomized, and the investigators were not blinded during experiments or outcome assessment.

### Study population and biospecimens

Male patients (approximately 35–75 years old) scheduled at our institution for a screening or surveillance colonoscopy were eligible for recruitment to MAGIC Biobank (https://www.magic.path.utah.edu/biobank) under IRB #00113193 through the University of Utah. Baseline characteristics of the study cohort are available in **Table 1**; we restricted the study to males because of sex-differences postulated in CRC. Peripheral blood mononuclear cells (PBMCs, cells isolated) were also collected from 19 of 21 patients for blood sequencing. Collaborating gastroenterologists collected colorectal biopsies using jumbo forceps from standardized regions including: cecum, ascending colon, transverse colon, descending colon, sigmoid colon, and rectum, as discernible by morphology during the procedure. Two endoscopic biopsies were taken from each region identified by the attending gastroenterologist. Colon biospecimens were immediately placed in 10% fetal bovine serum in sterile PBS for transport back to the laboratory. Within 30 minutes of acquisition, tissues were placed in OCT in a moulding block and solidified using dry ice. Samples were stored at -80°C until cryosectioning.

Cryomolds were cut into ∼15µm sections using a cryostat and mounted onto PEN slides to maximize crypt size along the longitudinal axis. A pilot cryosectioning was performed to ensure proper orientation of colon crypts (transverse “test tube” orientation). A dehydration mixture of 0.1 vol sodium acetate:2 vols of 100% ethanol was mixed and rested overnight to fix slides for 2 minutes prior to staining. H&E staining was performed on the Tissue-Tek® Prisma Automated Stainer (62667-SS) as follows: Wash station 1:00 (1 minute); Distilled water 0:10 (10 seconds); Hematoxylin 3:00; Wash Station 1:00; Ammonia water 1:00; Wash station 0:30; Eosin 1:30; Wash station 1:00. We proceeded with laser-capture microdissection within 24 hours of staining, as per protocol recommendations.^66^

### Laser-capture microdissection of colonic crypts

Laser capture to isolate individual colon crypts was performed using the MMI CellCut (102001), with contact capture into MMI provided isolation caps (product number 50204). The laser was routinely calibrated, and settings were consistently ∼78µm cut focus and 50-60% cut power, with capture at the objective 20x.

### DNA extraction and library preparation

Colon crypts were transferred from isolation caps with 20µl of reconstituted proteinase K solution (Arcturus: KIT0103) into eight-strip tubes compatible with thermocyclers. Visual confirmation of crypt transfer was possible in the majority of cases. Cell lysis was performed at 60°C (6 hours), denaturation at 75°C (30 min), and the samples were stored at 4°C. AMPURE bead purification (Beckman Coulter: A63880) was performed with two rounds of 75% ethanol washes; beads were kept in solute, and DNA quantification was not performed, as recommended to maximize DNA yield.^66^

PBMC samples for matched tissue germline filtering underwent DNA extraction via the Qiagen QIAamp Mini DNA Kit. High-molecular-weight DNA was fragmented using mechanical shearing to ∼350 bp (Covaris, Cat# 500295).

Low-input library preparation was performed for both crypt and blood samples (Watchmaker DNA Library Prep Kit part number 7K0101-096). We used IDT X-Gen UDI-UMI (part number 10005903) in the adapter ligation. Crypt samples underwent 8 cycles of PCR amplification. After library build, libraries were checked for size using the AATI Fragment Analyzer with the High Sensitivity NGS kit, and concentration was checked using the Qubit 1X dsDNA HS Assay Kit (part number Q33231). All samples were then converted using the Element Biosciences Adept Rapid PCR-Plus Protocol (kit 830-00018) with five PCR cycles.

### Whole-genome sequencing

Sequencing data (Element Biosciences AVITI and AVITI24) were generated according to the manufacturer’s recommendations with the Element Elevate library preparation kit (Element Biosciences, Cat# 830-00008).^40^ Linear libraries were quantified by quantitative PCR and sequenced with either the Cloudbreak Freestyle (n = 20, Element Biosciences, Cat# 810-00003) or Cloudbreak UltraQ (n = 28, Element Biosciences, Cat# 810-00008) sequencing kits.

Bases2Fastq Software (versions v2.1.0, v2.2.0, v2.2.1, and v2.3.0; Element Biosciences) was used to generate demultiplexed FASTQ files.

### Data processing and filtering

FASTQ files (R1 and R2) were aligned to the GRCh38 reference using BWA (0.7.19) with default parameters and 16 threads.

Bams underwent variant calling through two tools: the haplotype-based FreeBayes^67^ (1.3.4) and the deep-learning method DeepSomatic^68^ (1.9.0). FreeBayes variant calling was performed for each donor across 5 Mb regions of each chromosome, requiring 2 alternate alleles with a minimum qsum of 40. Donors PD34200 and PD34201 from Lee-Six et al. were omitted from genotyping for having 49 and 56 crypt samples, respectively, which was computationally prohibitive for freebayes’ donor-level joint calling. As both Lee-Six donors were healthy males with the study’s age range, we did not expect that these donors would provide additional biological context.

DeepSomatic variant calling was performed using containers downloaded from https://github.com/google/deepsomatic/blob/r1.9/docs/deepsomatic-quick-start.md, run on singularity^69^ (4.1.1) per chromosome at the sample level. Samples with matching blood were run with the “WGS” model type while samples without matching blood used the “WGS_TUMOR_ONLY” model type. Resultant sample- and chromosome-specific vcfs were then merged with GLNexus^70^ (v1.2.7) to produce per-donor results that were concatenated to comprise the entire genome.

The results of the variant callers were intersected to maximize stringency and ensure true biological signal in signature extraction.^71^ Any variant found in gnomAD^72^ v3 as annotated by Slivar^73^ was removed, as were low-complexity regions^74^ used for filtering in a previous lab study and loci from the UCSC genome browser^75^ Simple Repeats track, identified by Tandem Repeats Finder^76^. We kept non-reference mutations with zero reads in other donor samples (including blood), with a site depth of 8 and at least 4 alternate reads. An average depth of 10 was required to ensure consistent coverage across the genome. Variants found across different donors were removed as potentially missed population variants. Finally, samples with fewer than 500 single nucleotide variants (SNVs) were removed based on prior colon crypt work, presuming inadequate sample capture^11,53^, adjusted for this study’s distribution of SNV burden (**Supp. Fig. 1**).

### Signature analyses

Signature extraction was performed with SigProfilerExtractor^77^ (1.2.5), which is robust when compared with other extraction tools.^78^ The SBS288 schema, comprising stranded trinucleotide contexts, was used for SBS signature extraction, and the ID83 schema was used for insertion-deletion (indel) signature extraction. We used the reference genome GRCh38 and COSMIC version 3.4 for signature matching. A minimum of 1 and a maximum of 15 signatures was permitted in signature extraction, with default settings otherwise (non-negative matrix factorization replicates: 100, matrix normalization strategy: Gaussian mixture model; solution estimate stability: 0.8, minimum stability: 0.2, combined stability: 1.0). SBS signature extraction using all mutations had average stability of 0.96 and minimum stability of 0.9 (clonal: 0.94 and 0.86, subclonal: 0.99 and 0.99), and indel signature extraction of all mutations had average and minimum stabilities of 0.99 and 0.98 (clonal: 0.86 and 0.61, subclonal: 0.97 and 0.94). Mean correlation between original sample mutations and reconstructed signatures was ≥ 0.9 for all non-subclonal analyses, whereas the subclonal signature extractions had a mean correlation of 0.464 for the SBS mutations and 0.581 for indels.

Study-level, region-annotated signature information from Cornish et al. (2024) was taken from Supplementary Table 37. Patient-level, non-regional signature information from Diaz-Gay (2025) was taken from Supplementary Table 12; counts for microsatellite stable samples’ regions were derived from Supplementary Figure 9. We are unable to subset these data into clonal and subclonal mutations due to a lack of mutation allele frequency information. As CRC tumors contain a mixture of clonal and subclonal mutations,^79^ we elected to compare these tumor-specific data as-is to each signature extraction.

### Microsatellite analyses

We use the tool inSTRbility (GitHub: https://github.com/dashnowlab/inSTRbility) on all bams to evaluate microsatellite instability across samples.^80^ We use a catalog of five well-established microsatellite instability markers (BAT25, BAT26, NR21, NR27, NR24)^81^ and require two output instability indices reflective of mean and median allele size to be concordant (greater than 1 standard deviation above the average) to consider a locus unstable. If 1-2 loci were unstable, the sample was designated as MSI-L. No samples were 3+ unstable (MSI-H).

### Colibactin motif analyses

Huber et al. tested for motif enrichment in colibactin-exposed organoids compared to control organoids by recording all bases in a 10bp extended context around T>N mutations, which are associated with colibactin mutagenesis and in colibactin-associated mutational signatures SBS88 and ID18.^53^ The authors found that SNVs with adenine in the 3rd and 4th position of the sequence window were enriched after colibactin exposure, and were enriched most highly within 17 trinucleotide contexts showing significant cosine similarity to SBS88. Huber et al. further validated their colibactin-specific motifs using a random forest model trained on the organoids, and applied this pipeline to multiple cancer-sequencing datasets to identify colibactin-specific SNVs and indels. We used this pipeline to identify mutations in colibactin motifs in this study. All scripts used in analyses were directly adapted from the GitHub repository associated with Huber et al. (https://github.com/ProjectsVanBox/colibactin_detection).

### Statistical methods

We modeled data in the R statistical language (v4.5.2) with package lme4 (v1.1.38). We generally used general linear mixed models in the negative binomial space, as Poisson models were overdispersed. We included a random factor for donor identity and fixed factors for age—centered and scaled for model stability—and clinical group, labeled as cohort. The appropriate term for the level of anatomic detail—side (right and left), region (all six), or region_trinary (right, transverse colon, left)—was included as a fixed term in the various analyses. When incorporating Lee-Six data into these models, as when assessing the colibactin mutations across the three cohorts, we included the fixed factor of study, which was never significant. When including the GC content of callable bases in these models, the term was not significant and led to worse model fits (higher AIC and convergence issues), and was thus excluded. We also modeled a side*age test to assess differences in slope with age across the colon, but there was no significant interaction.

Our modeling based on count data (crypt-specific and colibactin motif SNVs and indels) used the log of the callable genome (number of bases in the reference genome with read depth ≥ 8) as an offset. For example, when modeling the accumulation of crypt-specific SNVs:

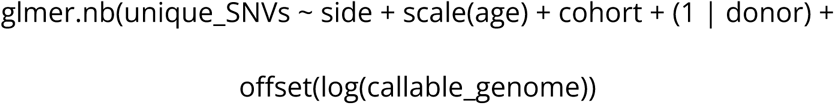

When modeling based on proportion, we used the log of the overall mutation count as an offset. For example, when modeling the C>[A,C,G] proportion significance across regions:

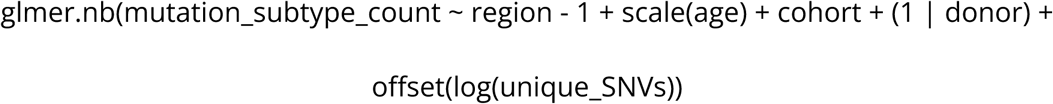

The term “region - 1”, which suppresses the intercept and instead uses a level mean rather than a reference level, was used in evaluating all regions for visualization in Figure 2, whereas “region” was used with apparent outlier rectum as the reference in determining statistical significance.

The exception to using negative binomial is the model of signature enrichment and depletion in regions. We used a logarithmic model to account for the binary absence or presence (region_bin) of a signature in a sample.

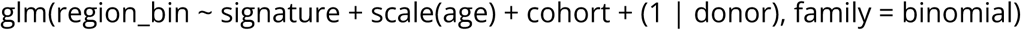

With regard to model stability, there were singular fits—indicative of likely overfitting—in the case of modeling mutation subtypes and signature enrichment/depletion, but we elected to keep the random effects variable (1 | donor) based on its probable biological significance. There was one instance of failed convergence when modeling the count of colibactin motif indels likely due to the lower counts in this scenario (occasionally less than 10, several fold lower than the colibactin motif SNVs), and so we evaluated the same model in a Poisson space with equivalent results (non-significance between clinical groups).

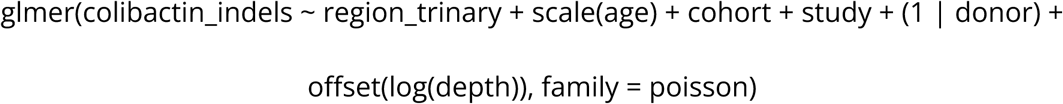

All models were corrected for non-independence by adjusting p-values with Benjamini-Hochberg correction.

### Data availability

Previously published whole-genome sequencing data were accessed from the European Genome-phenome Archive (EGA) with accession codes EGAD00001004192 through EDAM request 21757. Crams were downloaded and converted to fastqs using samtools and subsequently underwent identical processing to internally generated data.

Patient information for downstream analyses was derived from the Lee-Six et al. Supplementary Table 1 and the relevant Github repository (https://github.com/HLee-Six/colon_microbiopsies).

All software and code used are open source and freely available. Versions are documented within scripts, and all scripts, including the Snakemake pipeline with nested scripts, and produced data will be made available by publication.

## Data Availability

All data produced in the present study are available upon reasonable request to the authors and will be made available upon publication.

https://ega-archive.org/datasets/EGAD00001004192

https://github.com/HLee-Six/colon_microbiopsies

## Acknowledgments

The support and resources from the Center for High Performance Computing at the University of Utah are gratefully acknowledged. The computational resources used were partially funded by the NIH Shared Instrumentation Grant 1S10OD021644-01A1. Sequencing was supported by the University of Utah’s Center for Genomic Medicine. LH is supported by 1F30CA284847 from the National Cancer Institute.

LH thanks Kimberley Evason for providing initial histology expertise; Xiaoxu Yang for his leadership in bringing the MMI CellCut to the University of Utah and openness to sharing lab space; core specialist Ashley Vela and core director Derek Warner for their instrumental work and commitment to supporting this project; Allison Weis for clinical feedback; and Crystal Davey and Lilian Hayes of the Mutation Generation & Detection Core Facility for allowing LH to use their thermocycler. Additionally, the authors thank the gastroenterologists involved in sample collection—Kathryn Peterson, John D. Morris, Christopher Ko, John C. Fang, Jessica R. Stout, Andrew J. Gawron, Judith F. Staub, Matthew H. Steenblik, Darcie Gorman, Ann Flynn, Tuan Pham—as well as all patients who consented to be a part of our study.

## Author contributions

LH conceived the study, designed the experiments, and performed analyses with guidance from AQ and AMU. Initial biobank collection and sample processing, as well as documentation of patients’ clinical characteristics, were performed by EVP under AMU’s supervision. LH developed the molecular workflow with input and supervision from HH and structural support from JMH. LH developed the computational pipeline with bioinformatic support from JMM. AG supervised statistical methods development and provided essential insight in figure generation. CLG assisted LH with laser capture microdissection and DNA extraction. LH and AA modified the inSTRbility tool (originally written by AA) for study application to assess microsatellite instability, with clinical insight and supervision provided by DWN. TAS performed initial piloting of DeepSomatic variant calling and provided the codebase used in the pipeline. LH wrote the original manuscript and led its editing and revision, with input from all authors.

