## Supplementary Figures for "Colibactin-associated mutations in the human colon appear to reflect anatomy and early exposure, not oncogenesis"

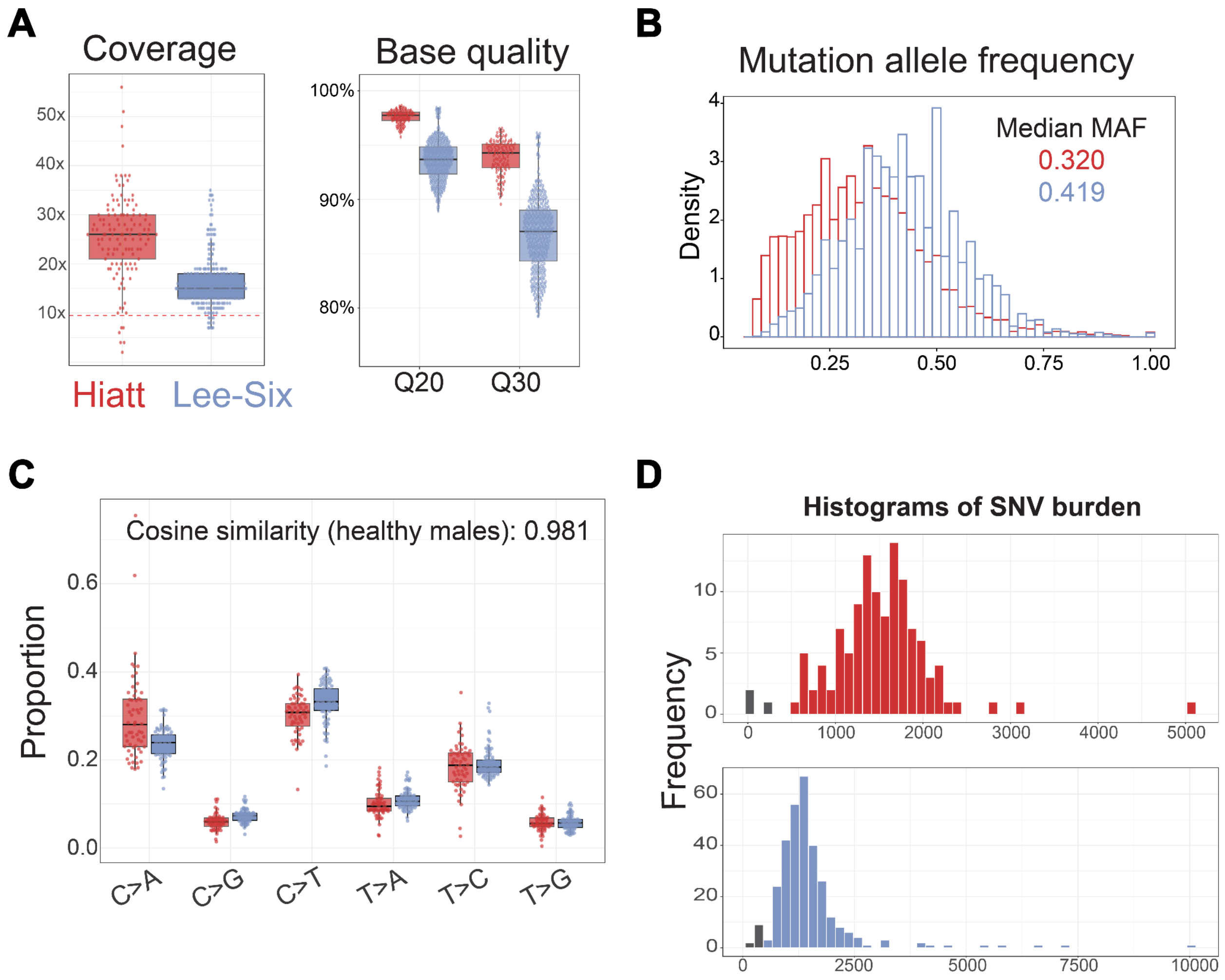


**Supplementary Figure 1: Overview of processed data from this study and Lee-Six et al. (2019).** Red is used for Hiatt data and blue for Lee-Six data. **A.** Coverage of both studies input from mosdepth reports, thresholded at 10x as used for analyses (Hiatt, n=118; Lee-Six, n=312), with a resulting median of 26x and 16x, respectively**.** Base quality from all Hiatt samples (n=145, blood samples included) and Lee-Six (n=445) taken from the fastp report. **B.** Mutation allele frequency (MAF) of all genotyped samples (Hiatt, n=125; Lee-Six, n=340) overlaid, with median MAF for each study group shown. **C.** Mutational proportion by six collapsed subtypes compared between healthy males in Hiatt (n=59) and Lee-Six cohorts (n=81), with associated cosine similarity of 0.981. Cosine similarity of full genotyped cohorts (Hiatt, n=125; Lee-Six, n=340) is 0.9694. **D.** Histogram of SNV burden between studies, with samples removed for having less than 500 crypt-specific SNVs in gray.

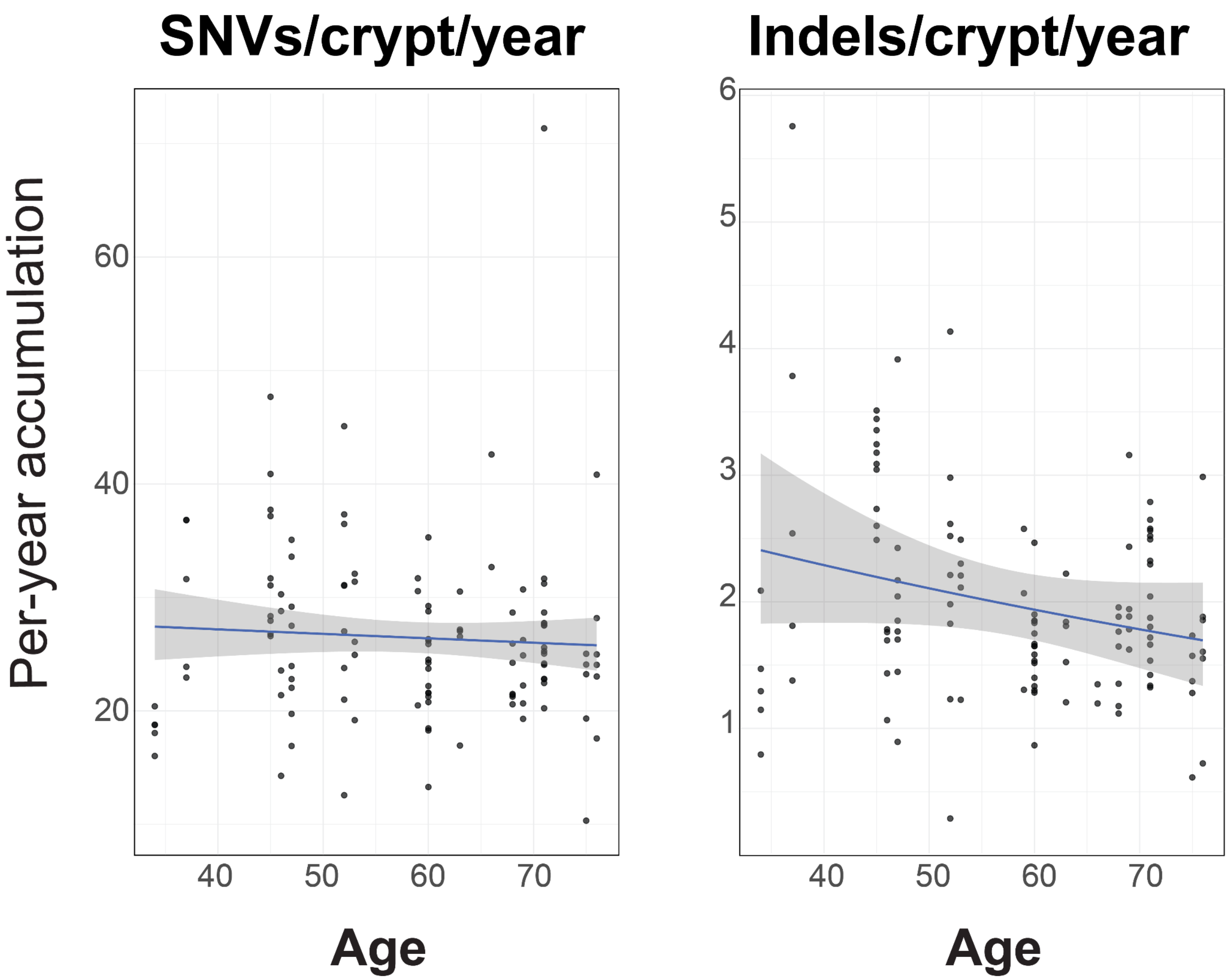


**Supplementary Figure 2: Per-year accumulation of crypt-specific mutations for each sample.** A negative binomial regression was performed for each mutation type, showing no apparent change over time in SNV accumulation and a potential decrease in indel accumulation over time.

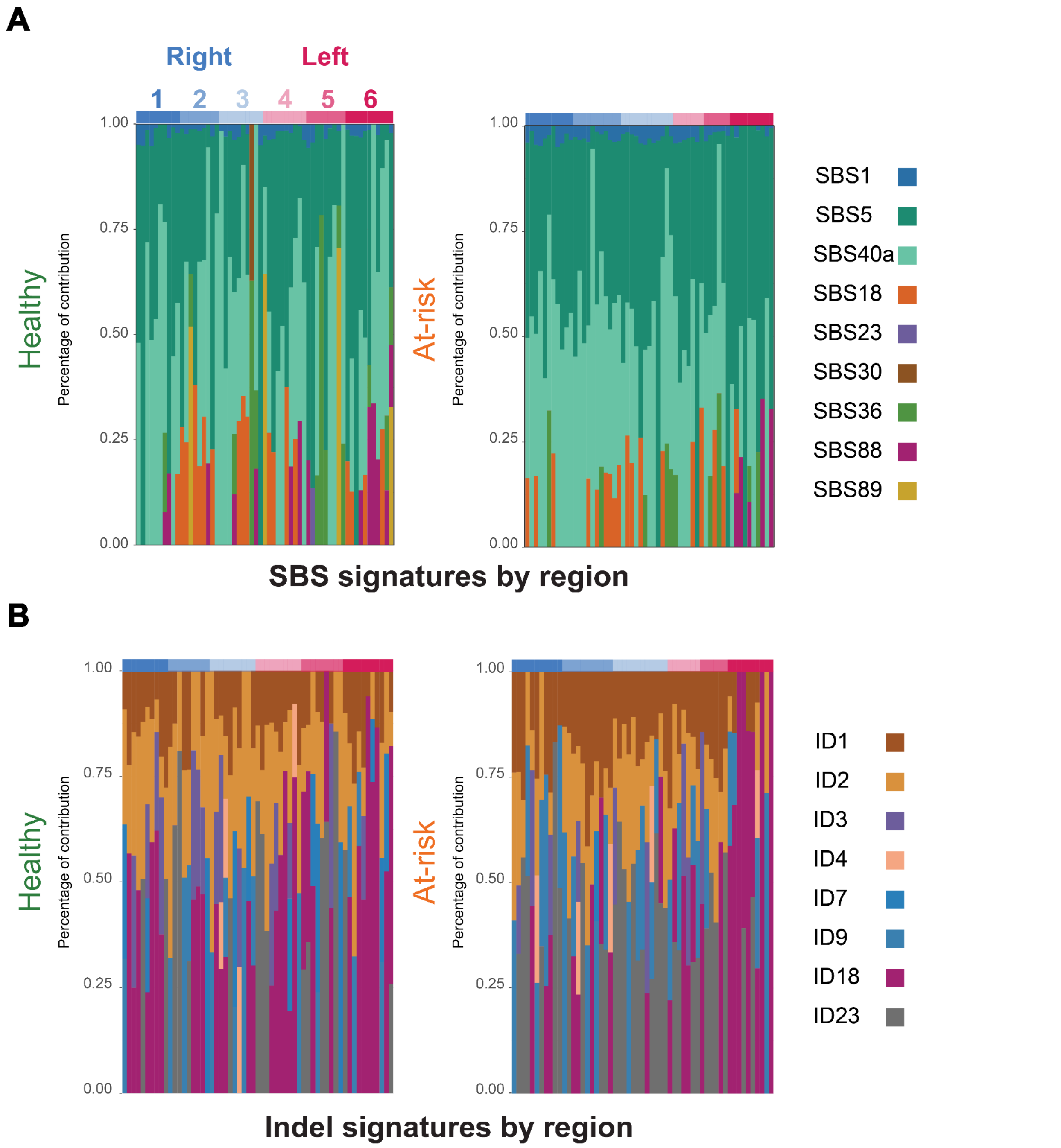


**Supplementary Figure 3: Signatures across regions, separated by clinical state.** Signature proportions of non-artifact signatures extracted across regions in premalignant groups. Samples include 59 healthy:57 at-risk samples, subset by region as 10:11 (cecum), 9:11 (asc. colon), 10:12 (trans. colon), 10:7 (desc. colonl), 9:6 (sig. colon), and 11:10 (rectum). **A.** SBS signatures **B.** Indel signatures.

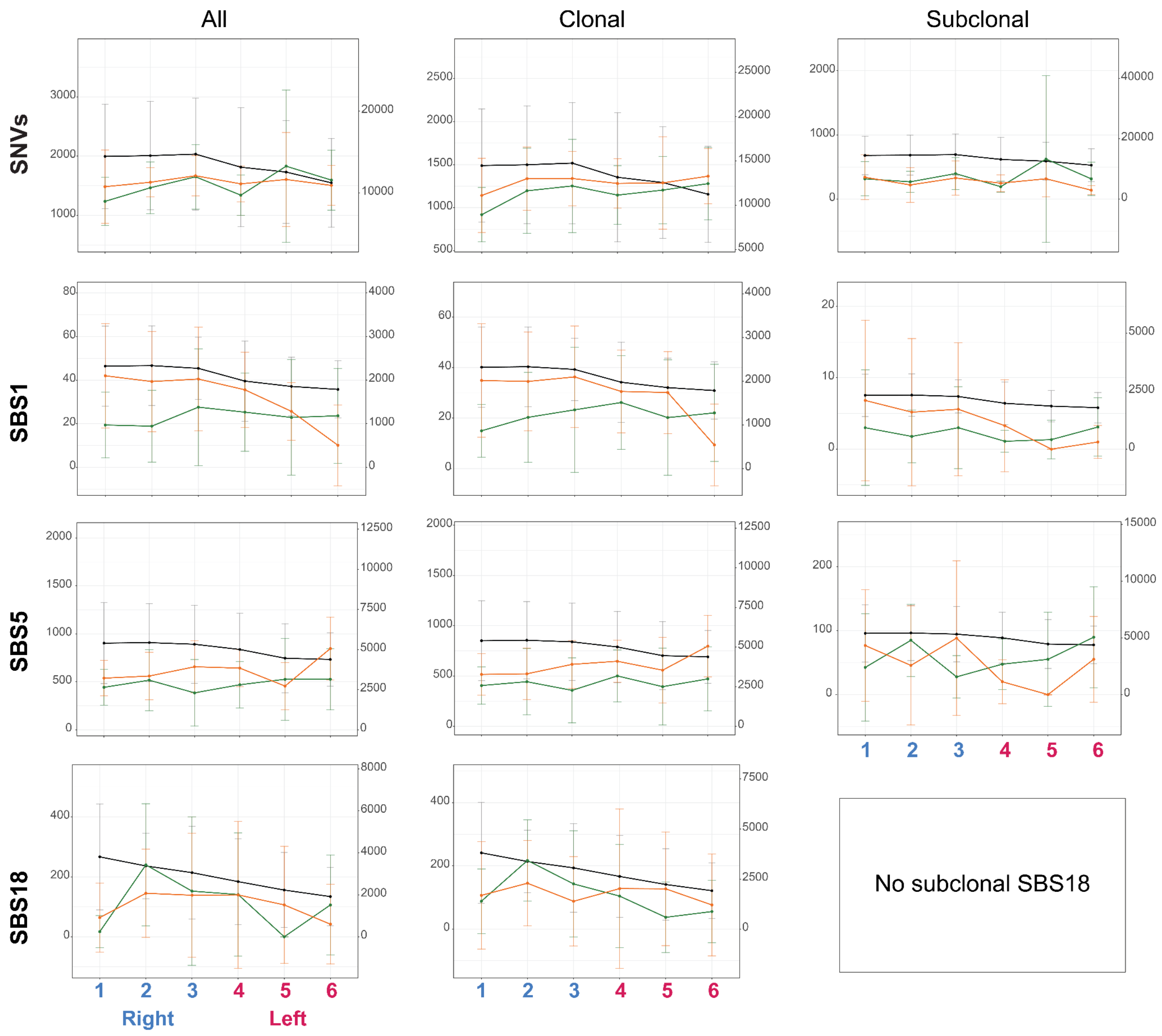


**Supplementary Figure 4: SNV burden and SBS signatures for three clinical groups.** Subset by MAF for clonal ($\geq$0.2) and subclonal (<0.2). Healthy: green, at-risk: orange, Cornish tumor trend: black. Mean and standard deviation are shown for all.

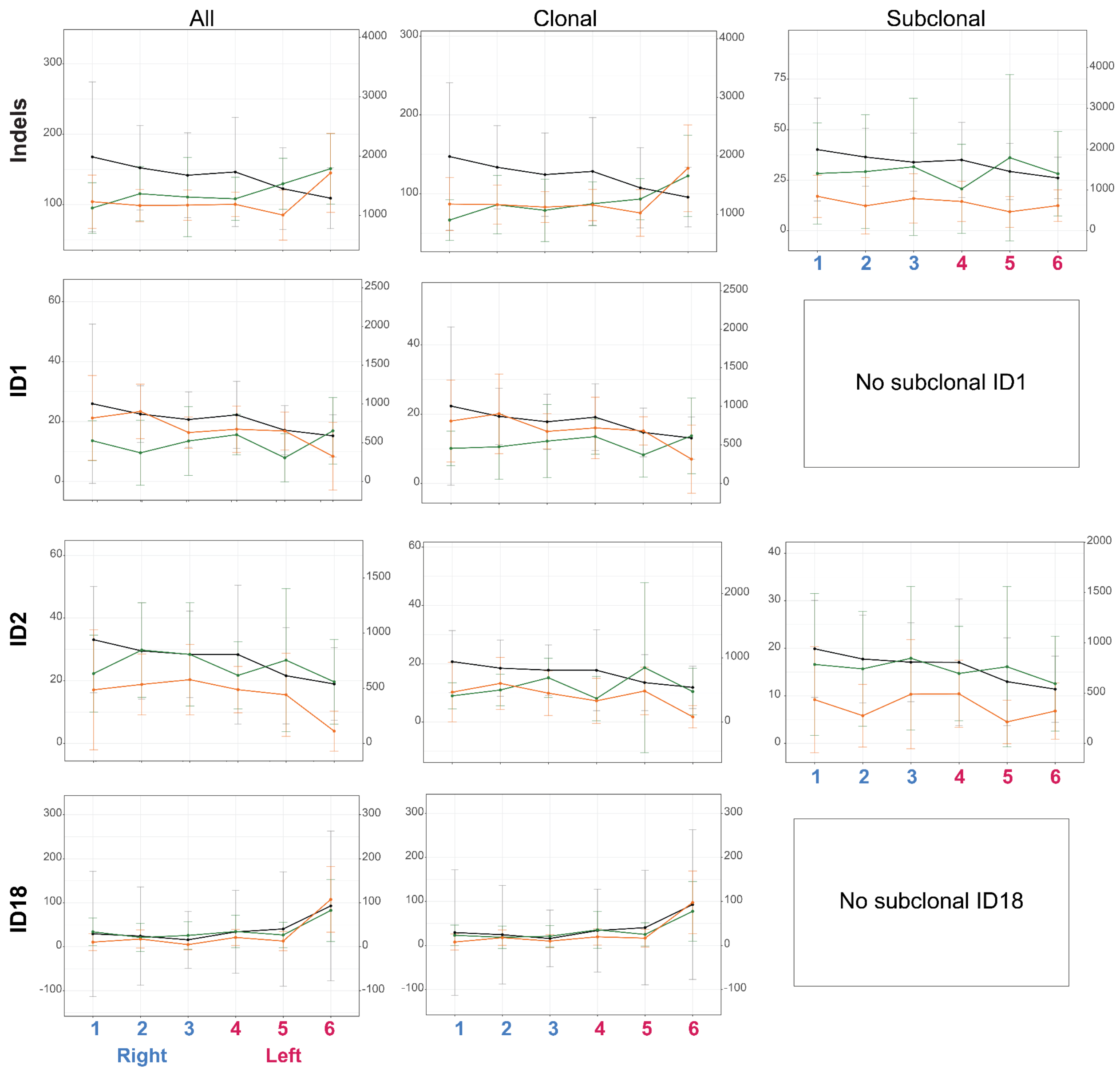


**Supplementary Figure 5: Indel burden and ID signatures for three clinical groups.** Subset by MAF for clonal (≥0.2) and subclonal (<0.2). Healthy: green, at-risk: orange, Cornish tumor trend: black. Mean and standard deviation are shown for all.

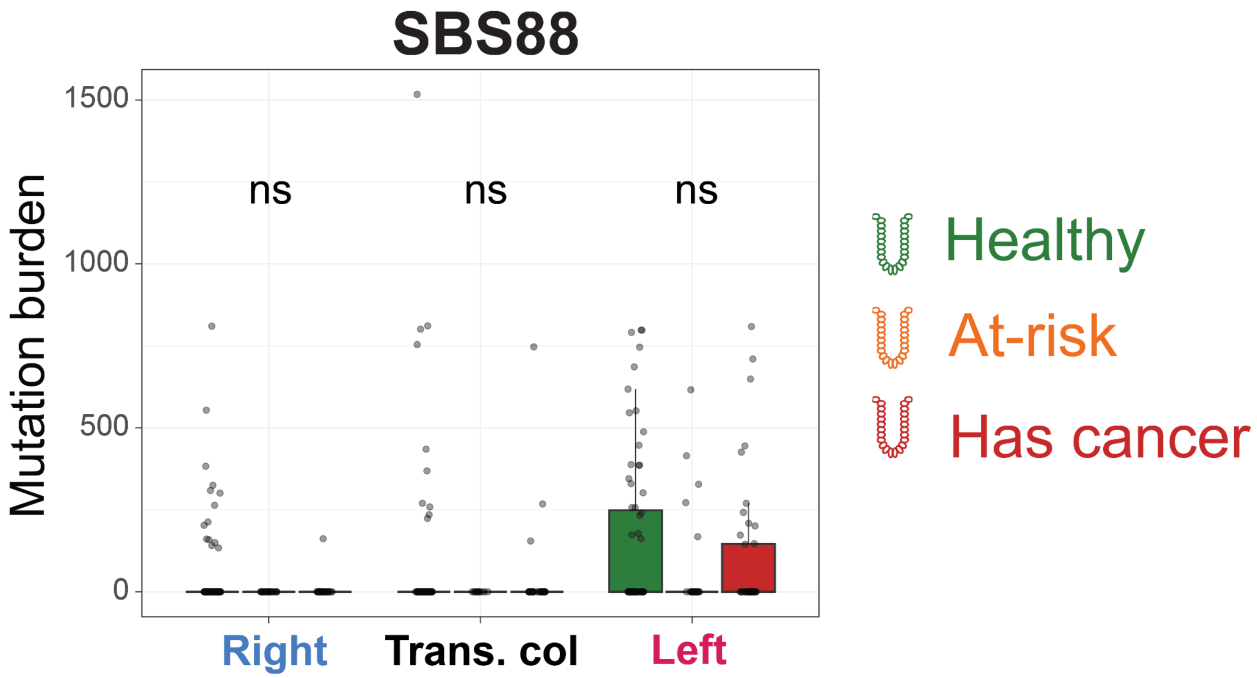


**Supplementary Figure 6: SBS88 by region across clinical groups.** Hiatt and Lee-Six et al. data included. Significance between clinical groups determined by GLM modeling with Benjamini-Hochberg correction (**Methods**).

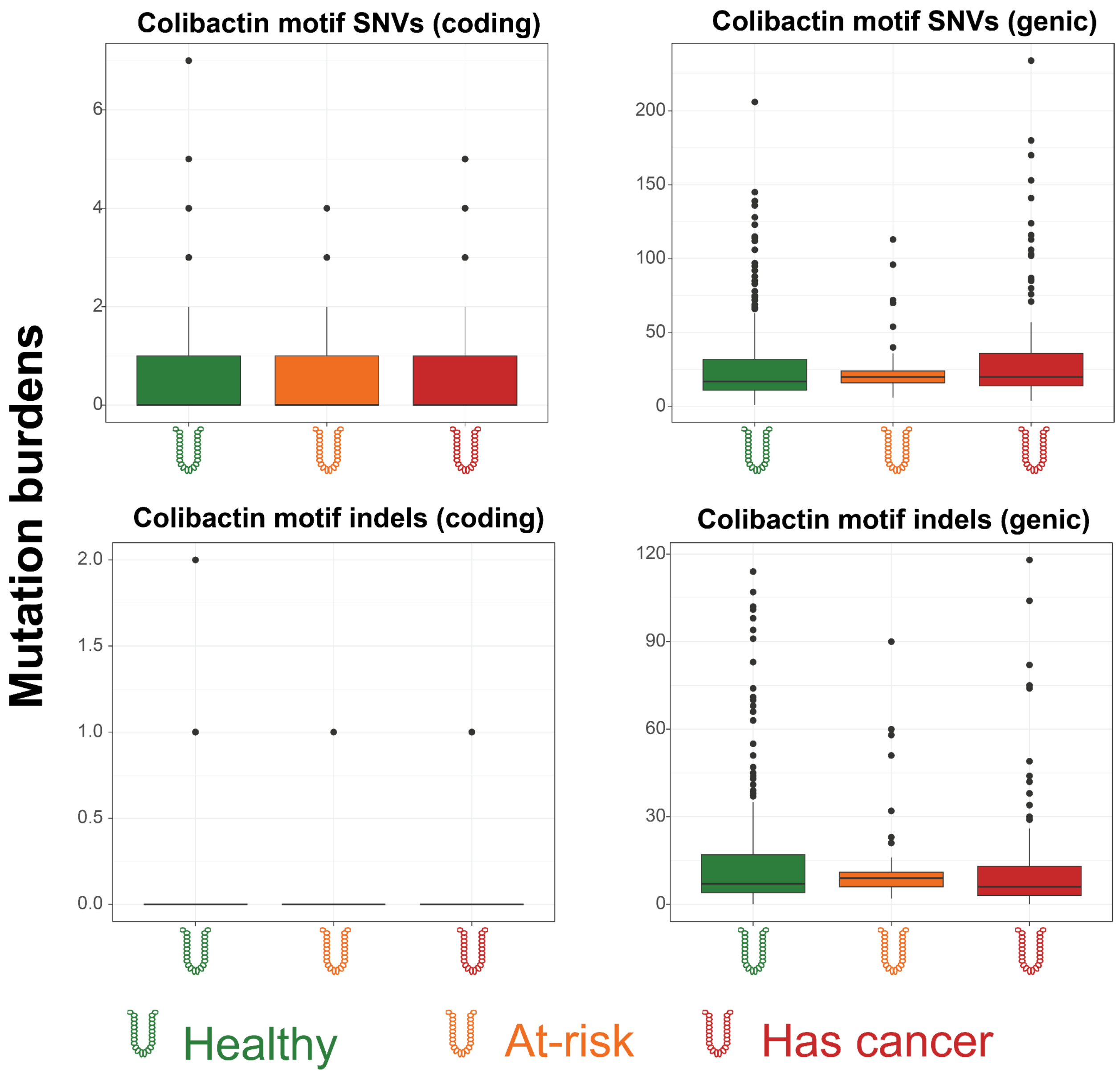


**Supplementary Figure 7: Comparison of colibactin motif mutation burdens across clinical groups.** Hiatt and Lee-Six et al. data included. Coding mutations are those found explicitly in exons, whereas genic mutations overlap with a gene. Kruskal-Wallis testing with Benjamini-Hochberg correction found no significant differences across these four cases; we did not use modeling here due to the extremely low mutation count, which led to convergence issues and other forms of model instability.

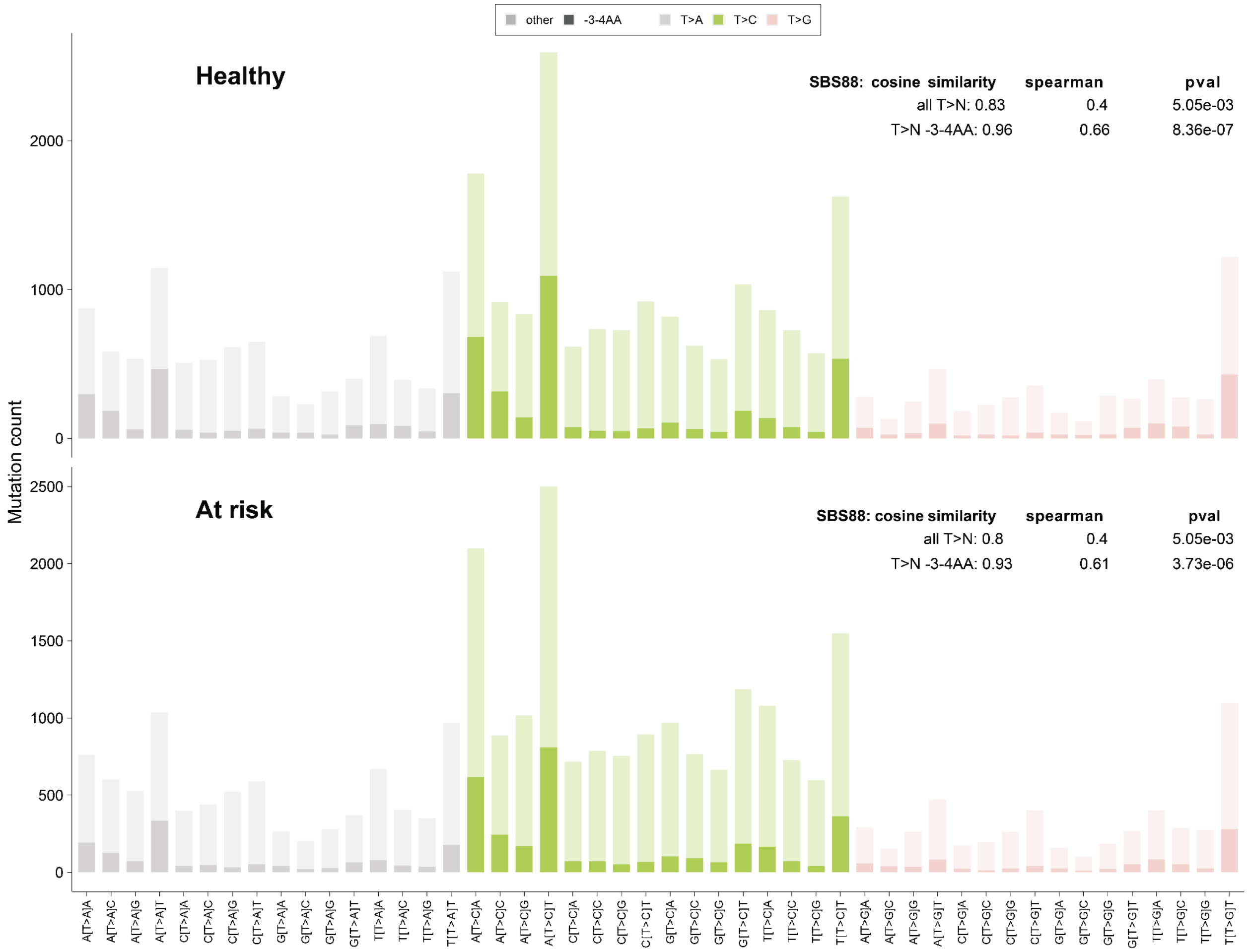


**Supplementary Figure 8: Cosine similarity of colibactin motif SNVs to SBS88.** Data restricted to Hiatt study. The colibactin motif uses T>[A,C,G] mutations with A nucleotides in the 3rd and 4th positions; all T>[A,C,G] mutations, regardless of motif, are compared to SBS88 for comparison. Spectra and statistics are separated into the two premalignant clinical groups: FDR-adjusted p-values are derived from a paired correlation test using Spearman's statistic.

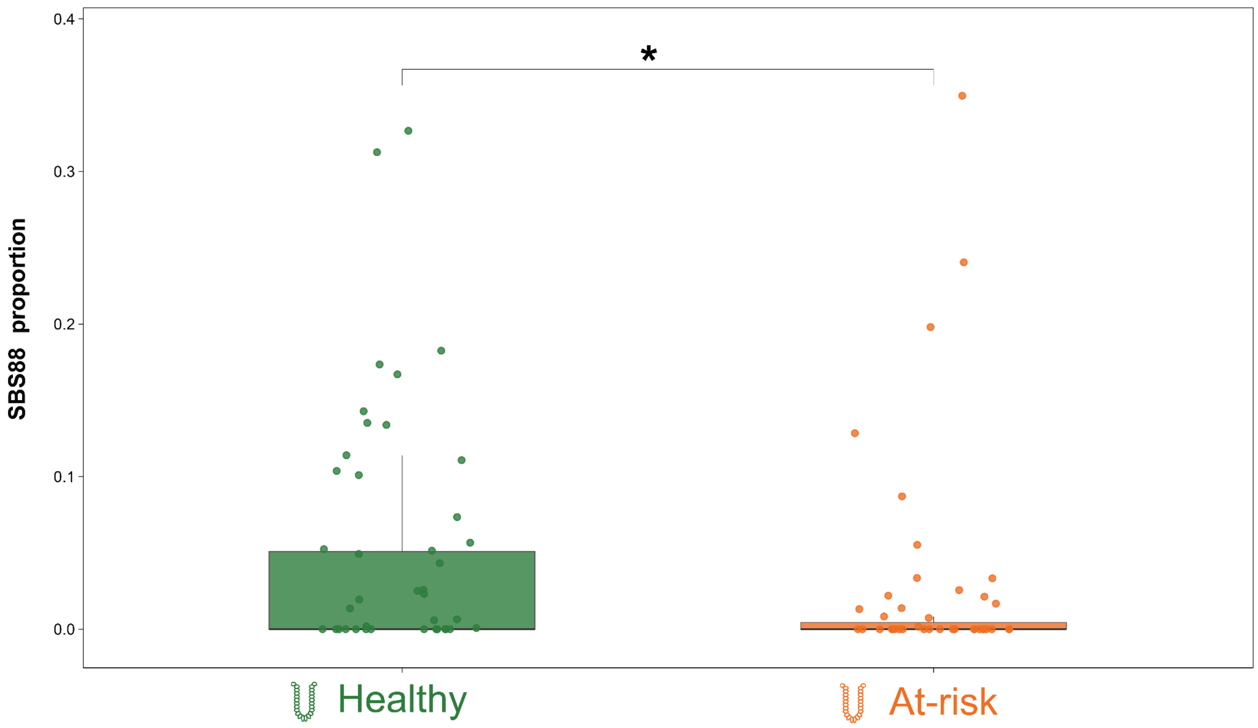


**Supplementary Figure 9: SBS88 proportion between clinical groups**. P-value (0.033) determined by Kruskal-Wallis test. Compared with Supplementary Figure 6, which shows that the raw mutation burden is consistent across groups, the SBS88 proportion differs significantly.

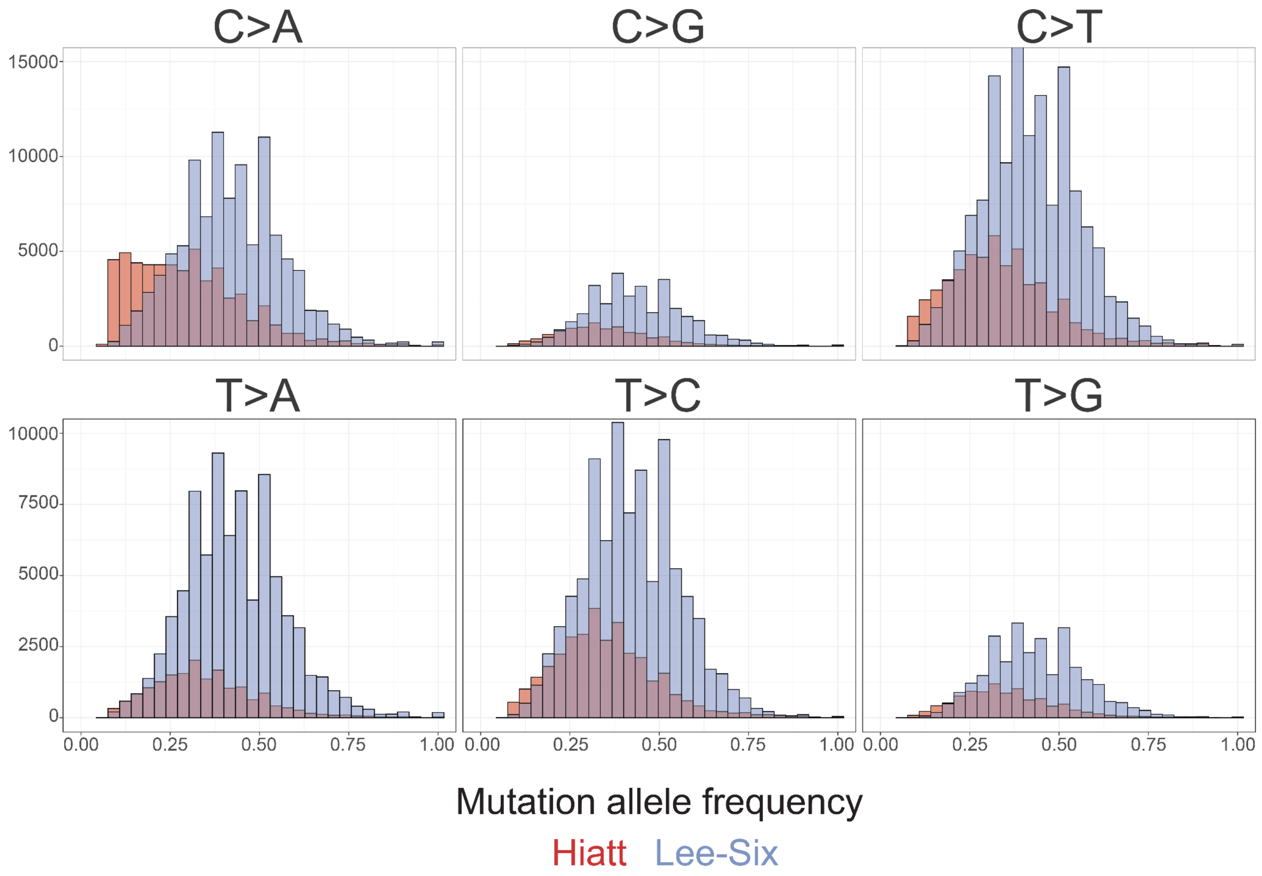


**Supplementary Figure 10: Mutation allele frequency by mutation subtype.** Hiatt is shown in red; Lee-Six data in blue. The mutation subtypes are broadly symmetric across studies, with the Hiatt MAF left-shifted due to increased subclonal capture. The exception is the C>A subtype, which shows elevated subclonal mutations that may reflect sporadic artifacts in the data.
