## Supplementary Tables for "Colibactin-associated mutations in the human colon appear to reflect anatomy and early exposure, not oncogenesis"

| **Overlapping signatures** | **Biological significance** | **Tumor trend** | **Match in regional trends?** | **Interpretation** |
|---|---|---|---|---|
| *SBS1* | Spontaneous deamination; clock-like | Decreased right-to-left | No; more comparable in at-risk clinical group | Hypermutability associated with microsatellite instability absent in our data; age or advanced clinical state may influence mutagenesis |
| *SBS5* | Clock-like; associated with cell division/aging | Decreased right-to-left | No | Hypermutability… |
| *SBS18* | Oxidative damage | Decreased right-to-left | **Yes** | Oxidative damage is elevated in the proximal colon |
| *ID1* | Slippage during replication; clock-like | Decreased right-to-left | No; more comparable in at-risk clinical group | Hypermutability… age or clinical state… |
| *ID2* | Slippage during replication; clock-like | Decreased right-to-left | Slightly**;** more comparable in at-risk clinical group *and* in subclonal mutations | Hypermutability… age or clinical state… |
| *ID18* | Colibactin | Increased right-to-left | **Yes** | Colibactin mutagenesis enriched in the distal/rectal colon |

**Supplementary Table 1: Contextualization of Cornish et al. signatures overlapping with this study.**

References associating hypermutability with microsatellite instability in the proximal colon:

1. (Farmanbar et al. 2023).
2. (Kim et al. 2024).
3. (Xu et al. 2024).
4. (Ward et al. 2025)

| **Anatomic term** | **Statistical Test** | **Mutation subtype** | **Adj. p-value (*p-val.*)** |
|---|---|---|---|
| Side (Right vs left) | Paired Wilcoxon test | C>A | 0.380 (*0.380)* |
| | | C>G | 0.051 (*0.014*) |
| | | ***C>T** | **0.027 (*0.003*)** |
| | | CpG>TpG | 0.083 (0.059) |
| | | T>A | 0.051 (*0.029*) |
| | | T>C | 0.051 (*0.029*) |
| | | T>G | 0.280 (*0.240*) |
| Region (Cecum, ascending colon, transverse colon, descending colon, sigmoid colon, rectum) | Kruskal-Wallis test | ****C>A** | **0.008 (*0.003*)** |
| | | ***C>G** | **0.042 (*0.030*)** |
| | | C>T | 0.081 (*0.070*) |
| | | CpG>TpG | 0.215 (*0.215*) |
| | | *****T>A** | **< 0.001 (< *0.001*)** |
| | | ****T>C** | **0.001 (< *0.001*)** |
| | | ***T>G** | **0.031 (*0.018*)** |

**Supplementary Table 2: Non-GLM statistical overview of mutation subtypes by location.** All p-values adjusted by Benjamini-Hochberg correction. Significant adjusted p-values are bolded alongside mutation subtype, with asterisks indicating significance level: *** < 0.001, ** < 0.01, * < 0.05.

| **Source** | **Study description** | **Included** | **Excluded** | **MAF?** |
|---|---|---|---|---|
| Our study | Whole-genome sequencing of 125 **“normal” crypts** from 21 individuals | **116** crypts (**21** patients) | Nine crypts<br>- 6 with coverage < 10<br>- 3 crypts with total SNV burden < 500 | Yes |
| Lee-Six et al., 2019 | Whole-genome sequencing of 445 **“normal” crypts** from 42 individuals<br>- | *Quality comparisons:* **445** crypts (**42** patients)<br><br>*Genotyped (340 total):* **301** crypts (**38** patients)<br>- 113 crypts from 15 patients with CRC<br>- 188 crypts from 23 healthy individuals | *Not genotyped:* 2 patients (105 crypts); too many samples to joint call with freebayes<br><br>*Genotyped (340 total):*<br>- 16 crypts removed for being ileum (small intestine)<br>- 12 crypts removed for having coverage < 10<br>- 11 crypts removed for having <500 crypt-specific SNVs | In genotyped data |
| Cornish et al., 2024 | Whole-genome sequencing of 2,023 colorectal cancer samples (**tumors**) from participants in the UK 100,000 Genomes Project | **766** regionally annotated tumors<br>- Cecum (n=158)<br>- Ascending colon (n=79)<br>- Transverse colon (n=60)<br>- Descending colon (n=34)<br>- Sigmoid colon (n=262)<br>- Rectum (n=173) | 140 tumors at locations not in our dataset<br>- Hepatic flexure (n=21)<br>- Splenic flexure (n=21)<br>- Rectosigmoid (n=98)<br><br>1,117 tumors without anatomic site annotated | No, public data only |
| Diaz-Gay et al., 2025 | Whole-genome sequencing of 981 colorectal cancers (**tumors**) from 11 countries on 4 continents | **802** microsatellite-stable tumors<br>- Proximal (n=189)<br>- Distal (n=298)<br>- Rectal (n=315) | 153 microsatellite-unstable tumors; 26 tumors otherwise classified | No, public data only |

**Supplementary Table 3: Data manifest for study sources used in manuscript.**
